# Heterogeneity of response to Early Start Denver Model: Identifying developmental trajectories and predictors of cognitive outcomes

**DOI:** 10.1101/2025.10.30.25339168

**Authors:** I. Ilaridou, N. Kojovic, T. Chataing, K. Latrèche, F. Journal, S. Akhavan, C. Sandini, M. Schaer

## Abstract

Autism is a highly heterogenous neurodevelopmental condition that can significantly compromise the quality of life. For children showing atypical trajectories of development in their early years, early and intensive intervention has shown promise in enhancing the development of language, cognition, and social skills. Yet, marked variability in intervention response remains insufficiently understood. Characterizing this heterogeneity is essential for informing intervention strategies tailored to each child’s developmental profile. The aim of the present work was to identify subgroups based on intervention response trajectories and investigate subgroup-specific predictors of intervention outcomes. We analyzed longitudinal data from 125 children (15.2 - 42.0 months) receiving the Early Start Denver Model (ESDM) intervention over an average follow-up period of 2 years (mean = 1.93, ± 0.25). Children gained an average of 17 points in Developmental Quotient (DQ), showed significant improvement in their adaptive skills and exhibited diminution of autism features. Using K-means clustering, we identified 3 subgroups: Progressive Group A (PrGA; 36%), which had the highest baseline cognitive scores (82.3) and gained 17 DQ points in average throughout the intervention; Progressive Group B (PrGB; 41.6%), which showed significant developmental delay at baseline (mean DQ = 56.3) and gained 30 DQ points; and the Persistent Support Needs Group (PSNG; 22.4%), which also started in the range of delay (mean DQ = 47.7) and remained in this range throughout intervention (mean DQ = 41.1). Subgroup-specific analysis revealed distinct patterns of baseline factors, including stronger adaptive skills and fewer autism features, associated with a higher cognitive outcome for PrGA and PrGB. Comparison of PrGB and PSNG further identified factors distinguishing children with similar levels of cognitive delay at baseline who nevertheless followed markedly different developmental trajectories. Lower levels of repetitive and restricted behaviors, stronger adaptive functioning, and greater cumulative intervention exposure characterized the PrGB group, suggesting that both child-related and intervention-related factors may contribute to variability in developmental outcomes.

**Lay Summary:**

- In a group of 125 autistic children receiving Early Start Denver Model intervention, we observed significant gain in cognitive and adaptive skills as well as reduced levels of autism features.
- We identified 3 subgroups of responses with different developmental trajectories. Distinct baseline factors were associated with the intervention outcome in different subgroups indicating that predictors of response are not the same for all children.
- These findings highlight the importance of personalized approaches to early intervention to enhance the developmental outcomes of autistic children.

## 1 INTRODUCTION

Autism is a complex neurodevelopmental condition characterized by social and communicational challenges as well as restricted and repetitive interests or behaviors (American Psychiatric Association, 2022). Early diagnosis of autism is essential as it enables timely access to early intervention designed to support developmental needs (Schreibman et al., 2015). Current literature suggests that early intensive intervention can benefit multiple developmental domains such as cognition, language and motor skills (Fuller & Kaiser, 2020; Virués-Ortega, 2010; Warren et al., 2011).

Among the various intervention approaches designed for young autistic children, the Early Start Denver Model (ESDM) is a well-established evidence-based intervention (Fuller et al., 2020). This approach aims to enhance skills in different developmental fields including receptive and expressive communication, social and cognitive skills, fine and gross motor as well as adaptive behavior (Rogers & Dawson, 2010). Since the initial Randomized Controlled Trial (RCT) study reporting significant cognitive gains in young children receiving 2 years of ESDM intervention relative to community-based services (Dawson et al., 2010), numerous studies have corroborated substantial improvements in cognitive and language outcomes associated with ESDM in larger samples (Godel et al., 2022; Mandelli et al., 2025; Rogers et al., 2019, 2021; Sterrett et al., 2025). Despite its highly standardized implementation, substantial variability in outcomes is consistently observed across children enrolled in ESDM. This variability highlights the heterogeneous nature of developmental responses among autistic children (Contaldo et al., 2020; Godel et al., 2022; Howlin et al., 2009; Sterrett et al., 2025).

Given this developmental heterogeneity, considerable effort has been devoted to identifying baseline characteristics associated with intervention response (Chetcuti et al., 2025; Panganiban et al., 2025; Vivanti et al., 2014). Cognitive abilities at the start of intervention, and particularly expressive and receptive language, have emerged as some of the most consistently reported predictors of favorable outcomes (Asta & Persico, 2022; Sinai-Gavrilov et al., 2020). Other predictors have also been proposed, including stronger adaptive communication skills (Godel et al., 2022) and eye-tracking–derived measures such as more social orienting or attention to faces (Latrèche et al., 2021; Robain et al., 2020). However, despite this body of research, findings across studies remain inconsistent. For instance, while some studies identify younger age at intake as a significant predictor of positive outcomes, others report no association with age (Chetcuti et al., 2025; Contaldo et al., 2020; Lombardo, 2021; Vivanti et al., 2016). These inconsistencies may stem from differences in study design across ESDM trials, site effects (Rogers et al., 2019) including variations in delivery format (individual vs. group-based), intervention duration and intensity, as well as substantial interindividual variability. Together, these factors likely hinder the identification of robust and generalizable predictors of intervention response (Chetcuti et al., 2025; Fuentes et al., 2021; Lombardo, 2021). One possible explanation for the limited reproducibility of predictive factors is that children receiving early intervention do not constitute a homogeneous population and may follow distinct developmental trajectories (Lord et al., 2015; Mandelli et al., 2025; Vivanti et al., 2014). Consequently, predictors identified at the population level may not apply equally across all children, potentially contributing to the limited reproducibility of predictor findings across studies (Vivanti et al., 2014).

In light of developmental profile and intervention response variabilities, identifying subtypes with distinct developmental trajectories is a critical step toward more personalized approaches to care (Chen et al., 2024; Journal et al., 2024; Kim et al., 2016; Latrèche et al., 2024; Nordahl et al., 2022). While a growing number of studies have applied data-driven stratification to observational autism cohorts, relatively few studies have examined the heterogeneity in the context of early intensive intervention, limiting our understanding of differential response to intervention. Asta and colleagues also attempted to disentangle heterogeneity by dividing a sample of 32 children who received nine months of ESDM into three subgroups (Asta et al., 2024). However, these subgroups were primarily defined by language skills and autistic features measured at the end of the intervention, and predictors within each subgroup were not examined. In a previous study involving 55 children receiving ESDM (Godel et al., 2022), we identified 3 subgroups with distinct trajectories of cognitive gain, however the limited sample size did not allow for the reliable identification of predictors of intervention related to cognitive gain. Examining subgroup-specific predictors of cognitive outcomes may provide valuable further insight into individual differences in intervention response. This strategy moves beyond the assumption that a single set of predictors applies to all children and instead acknowledges the variability in developmental trajectories. Critically, robust identification and validation of subgroup-specific predictors requires larger sample sizes than those used in previous.

In the present study, we leveraged a cohort of 125 children with a mean age of 29.8 months (15.2-42.0). Participants received ESDM intervention 12 to 20 hours per week over an average follow-up period of 2 years (mean = 1.93 ± 0.25) representing, to our knowledge, the largest monocentric sample assembled to further investigate the heterogeneity of response to ESDM. Our first aim was to evaluate the progress across the entire cohort in cognitive functioning as primary outcome, and adaptive skills and autism features as secondary. Our second aim was to identify baseline factors associated with the cognitive outcome by the end of the intervention. To achieve the latter aim, while also considering the multidimensional nature of autism and the interrelatedness of potential predictors, we adopted a multivariate approach using Partial Least Squares Correlation (PLS-C). The PLS-C identifies latent components that maximize covariance between two sets of variables. To this end, we examined whether age at intake, autistic features and adaptive behavior were related to the cognitive outcome at the last follow-up. Our third aim was to disentangle the heterogeneity in intervention responses by identifying subgroups of children with distinct developmental trajectories. To do so, we conducted a k-means clustering based on baseline cognitive scores and the rate of change of cognition throughout the intervention. We used PLS-C to identify whether the baseline factors examined above showed distinct patterns of association with developmental outcomes within each response group. Finally, to better understand the differences across the subgroups we applied a contrast PLS-C to compare their baseline characteristics and clinical measures, that potentially lead to distinct developmental trajectories.

## 2 METHODS

### 2.1 Participants

Since 2012, families enrolled in the ESDM program in Geneva were invited to take part in our study on developmental trajectories during intervention. The families were referred to ESDM because of clinically meaningful gaps in the child’s development. Participants received 12-20h per week of ESDM intervention in one of the Centres d’Intervention Précoce en Autisme (CIPA) in Geneva, Switzerland (see Godel et al., 2022 for a comprehensive description of the intervention program). Similarly to the original RCT (Dawson et al., 2010), the ESDM program is implemented for a duration of 2 years. Under specific circumstances, the duration of the program could be shorter (e.g. when the child reaches school age before the end of the 24 months period). We identified 135 autistic children who received ESDM intervention in the Geneva early intervention centers by Spring 2026 and had at least 1.5 years of follow-up. Nine were excluded due to incomplete data at baseline or at the end of the intervention and 1 participant was excluded due to a 6-month pause in the intervention. Our final sample thus consisted of 125 participants with an average age at intake of 29.8 months (± 4.92, see Table 1). From this final sample, 51 children were also included in our previous study that measured the outcome of ESDM after 2 years (of note, 4 children included in our previous study were not included in this study because of missing data from baseline evaluations necessary for the statistical analysis followed in this study).

**Table 1.**
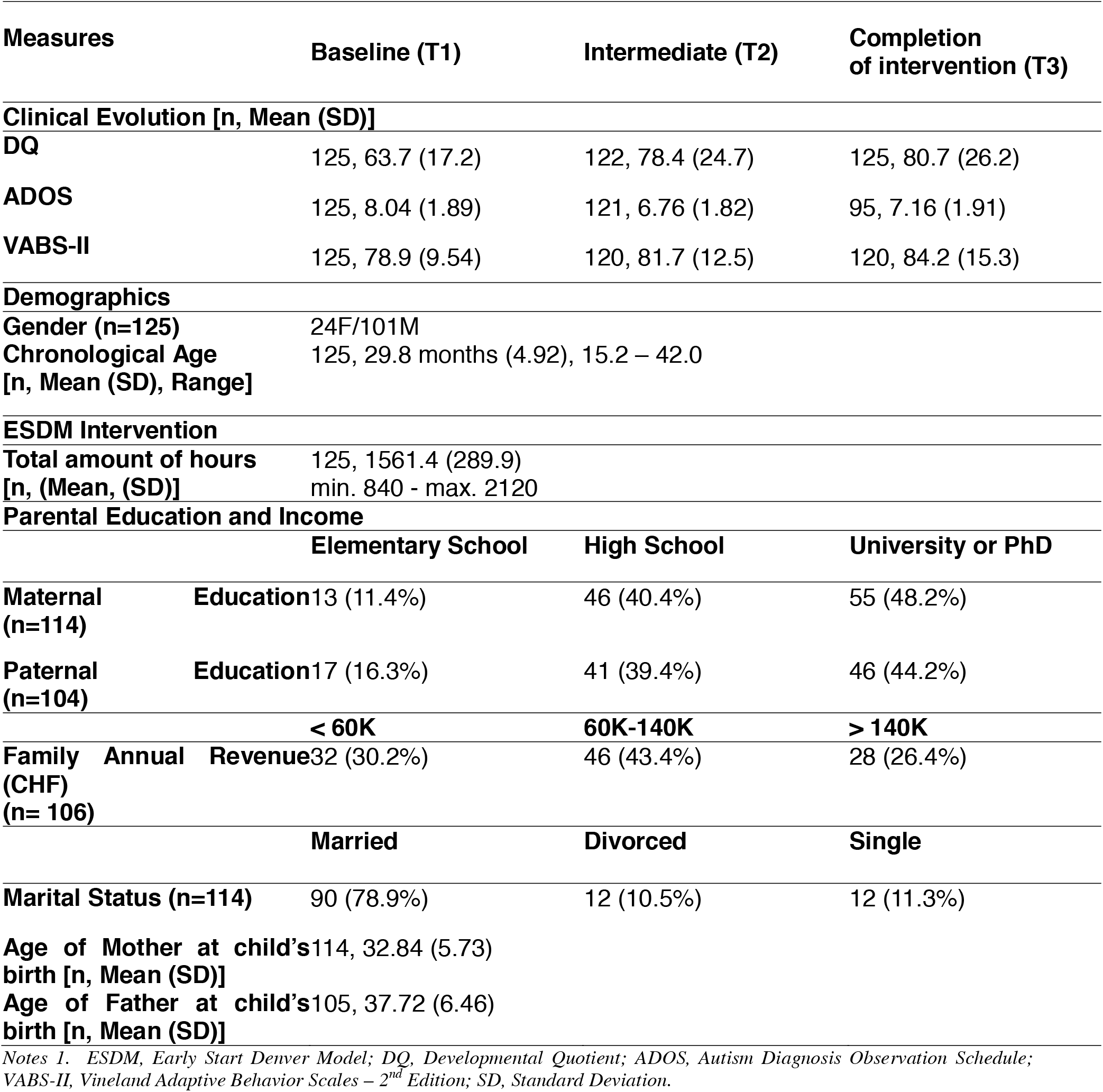
Sample characteristics throughout ESDM intervention.

All children received a clinical diagnosis of autism according to the criteria of the Diagnostic and Statistical Manual of mental disorders, 5th edition (American Psychiatric Association, 2013), which was corroborated with the gold standard diagnosis tool of Autism Diagnosis Observation Schedule-Generic (ADOS-G) (Lord et al., 2000) or 2nd Edition (ADOS-2) (Lord et al., 2012).

For the present study, the inclusion criteria consisted of evaluations of cognitive skills, ADOS and Vineland Adaptive Behavior Scales – 2^nd^ Edition (VABS-II) (Sparrow et al., 2005) at baseline (see more details in Measures section), a cognitive assessment at follow-up and completion of at least 18 months of intervention. For cognitive evaluation, we used the Mullen Scales of Early Learning (MSEL), the Psychoeducational Profile, Third Edition (PEP-3), and the Wechsler Preschool and Primary Scale of Intelligence, Fourth Edition (WPPSI-IV) (Mullen, 1995; Schopler et al., 2005; Wechsler, 2012). The choice of instrument depended on the participant’s date of inclusion, age, and level of language comprehension required to successfully complete the assessment. The MSEL was introduced into the protocol after 2015. At baseline, 23 participants were assessed with the PEP-3 instead of the MSEL; this was the case for 2 participants after one year and 3 participants at the end of the intervention. For 2 participants, only the WPPSI-IV was administered for the final cognitive evaluation due to families’ time constraints.

The 10 excluded participants did not differ significantly in cognitive skills (p = 0.892), adaptive skills (p = 0.597) or autism features (p = 0.446) at baseline from the 125 participants included in our analyses. Missing data were due to logistical issues (e.g., unavailable evaluation materials) or family-related difficulties leading to appointment cancellations or were conducted well before the start of the intervention and thus could not be considered true baseline measurements.

The research protocol has been approved by the Ethics Committee of the University of Geneva, and the caregivers of all the participants have signed an informed consent. Parents answered questionnaires about their child’s medical history and provided sociodemographic information (Table 1). Children were assessed at baseline after 6, 12, 18, and 24 months after the beginning of the intervention.

### 2.2 Measures

Autistic features were measured using the ADOS (Lord et al., 2012; Lord et al., 2000). The appropriate module was administered based on the child’s expressive language level and chronological age. To account for varying age and language levels, we used ADOS Comparison Score to quantify the level of autism features, as well as Social Affect (SA_ADOS_) and Restricted and Repetitive Behaviors (RRB_ADOS_) (Gotham et al., 2007). All ADOS assessments were administered by psychologists or medical doctors trained in ADOS administration. Assessments were videotaped, and later rated in team consensus with at least one examiner who had established research reliability on the ADOS-2. Research reliability was established following standard procedures, requiring at least 80% agreement with ratings provided by a certified ADOS trainer.

Similarly, participants underwent cognitive assessments with one or more of the following instruments: MSEL, PEP-3 and WPPSI-IV (Mullen, 1995; Schopler et al., 2005; Wechsler, 2012). To derive a reliable estimate of each child’s overall developmental level, we created a best-estimate developmental composite score following others (Godel et al., 2022; Howlin et al., 2014; Kojovic et al., 2019). The composite was constructed by selecting the most frequently used developmental assessment available for each child at each time point. When multiple assessments were available, we prioritized those with the highest representation across the sample (MSEL > PEP-3 > WPPSI-IV) to ensure consistency and comparability. Scores from the MSEL and PEP-3 were converted into a common metric - developmental quotient (DQ) - to facilitate integration across tools, and WPPSI-IV full scale IQ was used as equivalent (more details on supplement A). Since DQ is normalized for age and the timing of the evaluation, a decline in DQ does not necessarily indicate loss of skills but rather a slower rate of skill acquisition in the child compared to typical development. Additionally, a stable DQ score does not imply a lack of progress, but may reflect continuous development.

Finally, we used the VABS-II interview format to evaluate adaptive functioning across four domains: Communication (Com_VABS_), Daily Living Skills (DLS_VABS_), Socialization (Soc_VABS_), and Motor Skills (Mot_VABS_). We selected standard scores and the composite score representing overall adaptive behavior (Sparrow et al., 2005).

The ADOS, VABS-II, PEP-3 and MSEL were conducted at baseline, after 1 year and after 2 years of intervention.

### 2.3 Statistical Analysis

#### 2.3.1 General Trajectory of the sample

Statistical analyses and plots were performed using MATLAB R2024b, R version 2024 and IBM SPSS Statistics Version 29.0.2.0.

To evaluate the overall effects of the ESDM intervention, we examined changes in participants’ cognitive skills, adaptive functioning, and quantity of autism features at baseline (T1), after 1 year (T2) and after 2 years of intervention (T3) (See more details in Supplement A). Specifically, we conducted repeated measures ANOVA or, when the assumptions like normality or sphericity were violated, Friedman Test at the three timepoints (T1, T2, T3).

#### 2.3.2 Characterizing the distinct developmental trajectories

To investigate individual differences in response to the intervention, we applied the clustering approach previously described by Godel et al., 2022. First, we computed the rate of change by following the Symmetrized Percent Change formula:

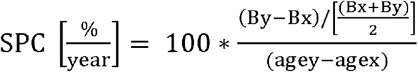

To identify the distinct developmental trajectories, we performed K-means clustering using DQ at T1 and the SPC of DQ from the onset until the end of the follow-up. The reasons behind the choice of cognition as the main intervention outcome and clustering variable are provided in Supplement A. Optimal number of clusters was determined using the SPSS in-built Two-Steps clustering algorithm with the log-likelihood as the distance measure and the Bayesian Information Criterion (BIC) as the clustering criterion (Chiu et al., 2001). This procedure identified a three-cluster solution, with a silhouette measure of cohesion and separation of 0.60, indicating good cluster quality.

In the next steps, we applied Partial Least Squares Correlation (PLS-C), a multivariate, data-driven statistical approach that identifies latent components representing patterns of maximal covariance between two sets of variables (Krishnan et al., 2011; McIntosh & Lobaugh, 2004). In the present study, PLS-C was used to examine whether multivariate profiles of baseline characteristics were associated with later cognitive outcome. Specifically, baseline measures included age at intake, autism features (SA_ADOS_ and RRB_ADOS_), and adaptive functioning domains from the VABS (Com_VABS_, DLS_VABS_, Soc_VABS_, Mot_VABS_), while final developmental quotient (DQ) served as the cognitive outcome measure. We first performed behavioral PLS-C including the overall sample and then separately within each subgroup to characterize subgroup-specific covariance patterns between baseline characteristics and later DQ. We then conducted a contrast PLS-C analysis to directly compare these covariance patterns between two specific subgroups. Statistical significance of latent variables was assessed using 1,000 permutation tests, and the stability of variable contributions was evaluated using 10,000 bootstrap resamples. Analyses were conducted using the publicly available myPLS MATLAB toolbox: https://github.com/MIPLabCH/myPLS.

#### 2.3.3 A3D Theoretical Framework

Among the alternative classification methods suggested in current literature is the A3D Theoretical framework, which distinguishes two subgroups, Type I and II, based on non-core features (language, cognitive, motor, and adaptive skills) rather than autism severity per se (Mandelli et al., 2024). Type I represents a slower and less pronounced improvement in development over time, with greater challenges and delays in skill acquisition. In contrast, Type II reflects a more typical development with milder difficulties and faster rate of developing skills compared to Type I. Since the A3D Theoretical framework provides an online app (https://land.iit.it/tools/a3dmselvabs/), we compared the outcome of the framework’s classification with ours.

## 3. RESULTS

### 3.1 General Trajectory of the sample

A description of the sample’s characteristics, including demographic variables, is presented in Table 1.

To assess the evolution of the sample over the course of the ESDM intervention, we compared the scores of Composite DQ, ADOS, and VABS-II across different time points. Because assumptions of normality and sphericity were violated, we conducted Friedman and Wilcoxon Signed-Rank Tests to compare related groups. Our findings indicate a statistically significant average increase of 17 points in Composite DQ, with more than 14 points gained during the first year of intervention (see Table 1 & Figure 1a). We also observed a statistically significant increase in adaptive behavior (Figure 1c) and reduction in the overall severity of autistic features across all timepoints (see Figure 1b). Precisely, across T1 and T3 we observed a significant decrease in the scores of SA_ADOS_ (p < 0.001) and a significant increase in the domain of RRB_ADOS_ (p = 0.014). More details about the trajectories of SA_ADOS_ and RRB_ADOS_ are provided in the Supplement B. The improvement in the SA_ADOS_ can be explained by the fact that ESDM primarily targets social skills such as communication, language and joint attention, leading to significant progress of SA_ADOS_ scores. In contrast, increases in RRBs dung the preschool years have previously been described in literature (Lord, 1995; Moore & Goodson, 2003) and given that RRBs are not the primary intervention target, a relative increase in this domain can be expected (Dawson et al., 2010; Schreibman et al., 2015).

**Figure 1.**
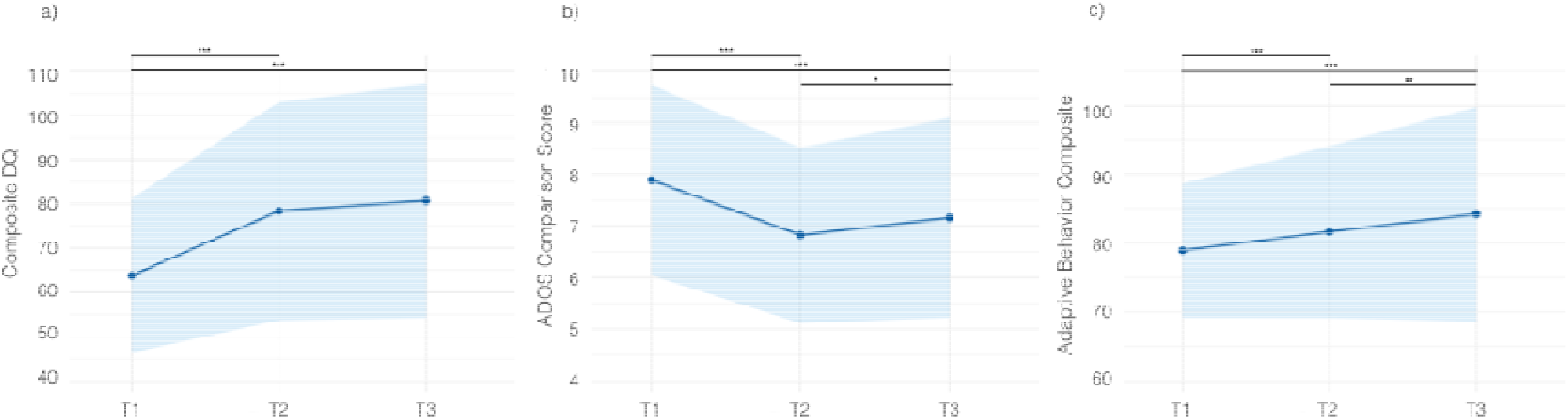
Evolution of participants throughout ESDM intervention. Notes. 1 Notes 2 Developmental trajectories of participants across: A) Composite DQ, B) ADOS Comparison Score, C) Adaptive Behavior Composite – Standard Score (VABS-II). Blue bands signifying Standard Deviation. Pairwise comparisons were conducted using Wilcoxon Signed Ranks Test. FDR applied for multiple comparisons. Each line chart was created including the participants without missing values across all three timepoints. The corresponding p-values were computed including participants without missing values between the timepoints of comparison. Scores with a significant statistical difference are highlighted in bold; *p < 0.05, **p < 0.01, ***p < 0.001. DQ, Developmental Quotient; ADOS, Autism Diagnosis Observation Schedule; Adaptive Behavior Composite; Composite score of Vineland Adaptive Behavior Scales – 2nd Edition.

### 3.2 Identifying heterogeneity of responses

According to the results of K-means clustering, we obtained 3 subgroups with distinct developmental trajectories. To further illustrate their differences at T1, Figure 2a-I presents violin plots comparing the 3 groups across baseline factors, including age at T1, DQ, subdomains of ADOS and VABS-II and SPC of DQ. Their clinical evolution and demographic characteristics are presented in Table 2 and Figure 2. Specifically, the first subgroup demonstrates a relatively higher Composite DQ, higher adaptive skills, and lower presence of autistic features at baseline. Moreover, it demonstrates marked progress across the ESDM intervention exhibiting important cognitive advancement compared to 2 other subgroups. Consequently, we refer to this subgroup as Progressive Group A (PrGA). The second subgroup follows a similar trajectory with PrGA. However, while the children of this group start with a significant developmental delay at baseline, they show substantial developmental gains, narrowing the gap with typically developing children and reaching the lower range of typical DQ development by the end of intervention. We denote this subgroup as Progressive Group B (PrGB). Finally, the third subgroup shows a significantly lower composite DQ, lower adaptive skill levels and higher autism feature levels at baseline compared to the other groups, and follows a rather stable trajectory of DQ scores. As mentioned in Methods, this stability in age-normalized DQ scores does not imply lack of progression or regression, but rather that this subgroup is not narrowing the developmental gap with their TD peers over time. Here, we observed that the average age equivalent scores increased by 7.87 points (±3.99) from T1 to T3. By the end of the intervention, this third subgroup has cognitive and adaptive skills that remain in the range of developmental delay indicating the need of further support. Thus, we decided to refer to this subgroup as Persistent Support Needs Group (PSNG).

**Figure 2.**
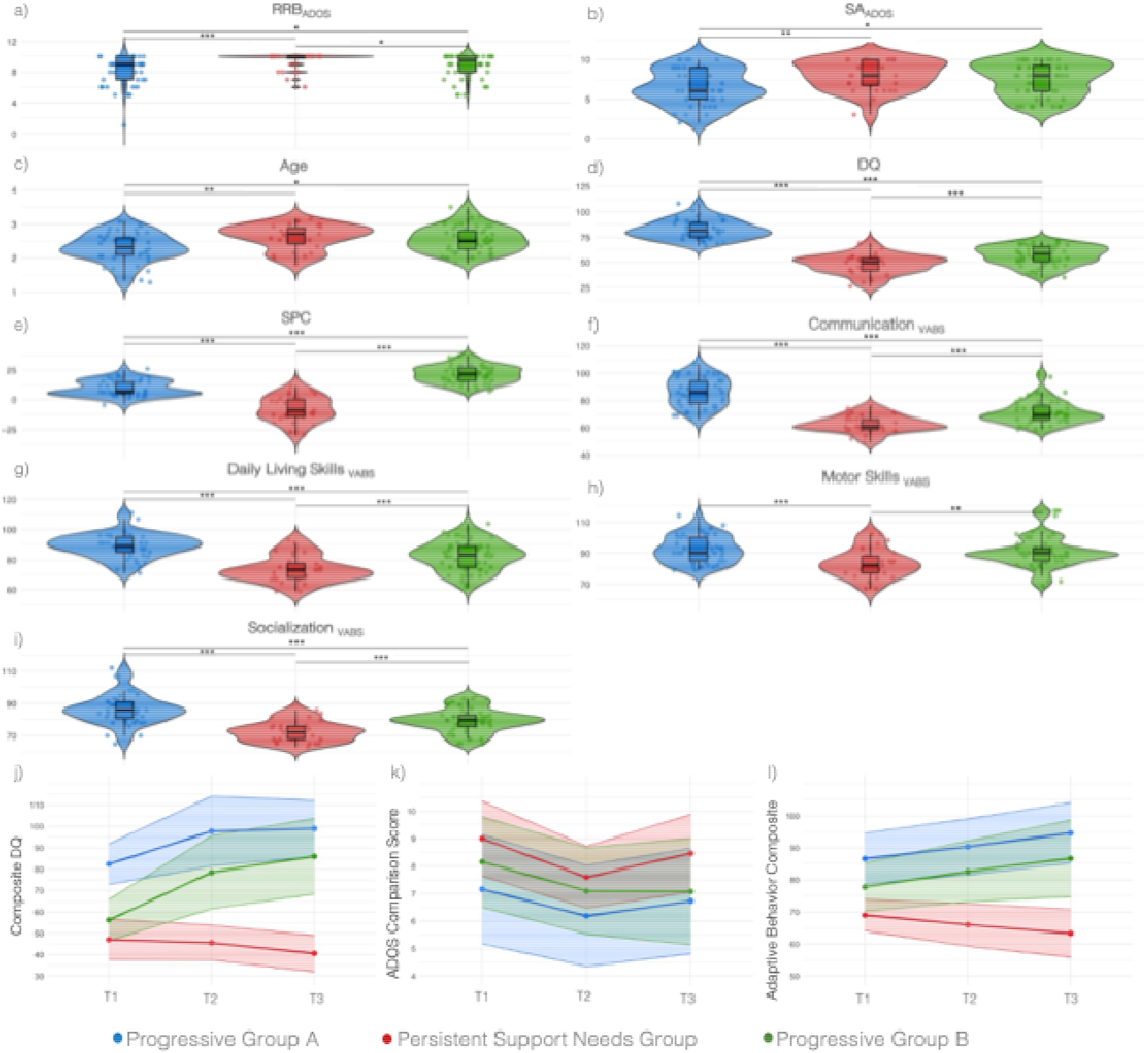
Comparison and developmental trajectories of the 3 subgroups. Notes. 2 Violin plots (a-g) with pairwise comparisons of the groups using Mann-Whitney U Test. FDR applied for multiple comparisons. Scores with a significant statistical difference are highlighted in bold; *p < 0.05, **p < 0.01, ***p < 0.001. Exact p-values are available in Supplement B, Table 2. Line charts (j-l) showing the developmental trajectories of the 3 subgroups across the 3 timepoints. Each line chart was created including the participants without missing values across all three timepoints. ADOS, Autism Diagnosis Observation Schedule; VABS-II, Vineland Adaptive Behavior Scales – 2^nd^ Edition; SA_ADOS,_ Social Affect; RRB_ADOS_, Restricted and Repetitive Behaviors; SPC, Symmetrized Percentage Change

Regarding the longitudinal trajectory of autism features, we observed a modest increase in ADOS total severity scores between T2 and T3 in both the overall sample and the response groups (Figures 1 and 2). In the progressive groups, this increase coincided with changes in the distribution of ADOS modules administered across timepoints. Specifically, a greater proportion of participants were evaluated using Modules 2 and 3 at T3, whereas assessments at earlier timepoints were predominantly conducted using the Toddler Module or Module 1. As Modules 2 and 3 involve more advanced social-communicative demands, this shift in module distribution may have contributed to the higher ADOS scores observed at T3. In contrast, within PSNG, most participants who showed higher ADOS scores at T3 were assessed using the same module (primarily Module 1) at both timepoints, suggesting that module transition alone does not fully account for the observed increase in this subgroup.

### 3.3 Predictors of the Cognitive Outcomes for the Original Sample

The next step was to identify which baseline variables were the most influential at predicting a higher cognitive outcome for the overall sample. According to the results of PLS-C, younger age, lower autism feature levels (SA_ADOS_ and RRB_ADOS_) and higher scores in all the domains of adaptive skills at baseline were associated with a higher final cognitive outcome (p = 0.001, r = 0.69) (Figure 3.1a). Additional analyses including the total number of intervention hours and demographic variables, such as parental age at the birth of the child, parental education, and family income, did not identify these factors as stable contributors to the latent component associated with cognitive outcome at the end of the intervention.

**Figure 3.**
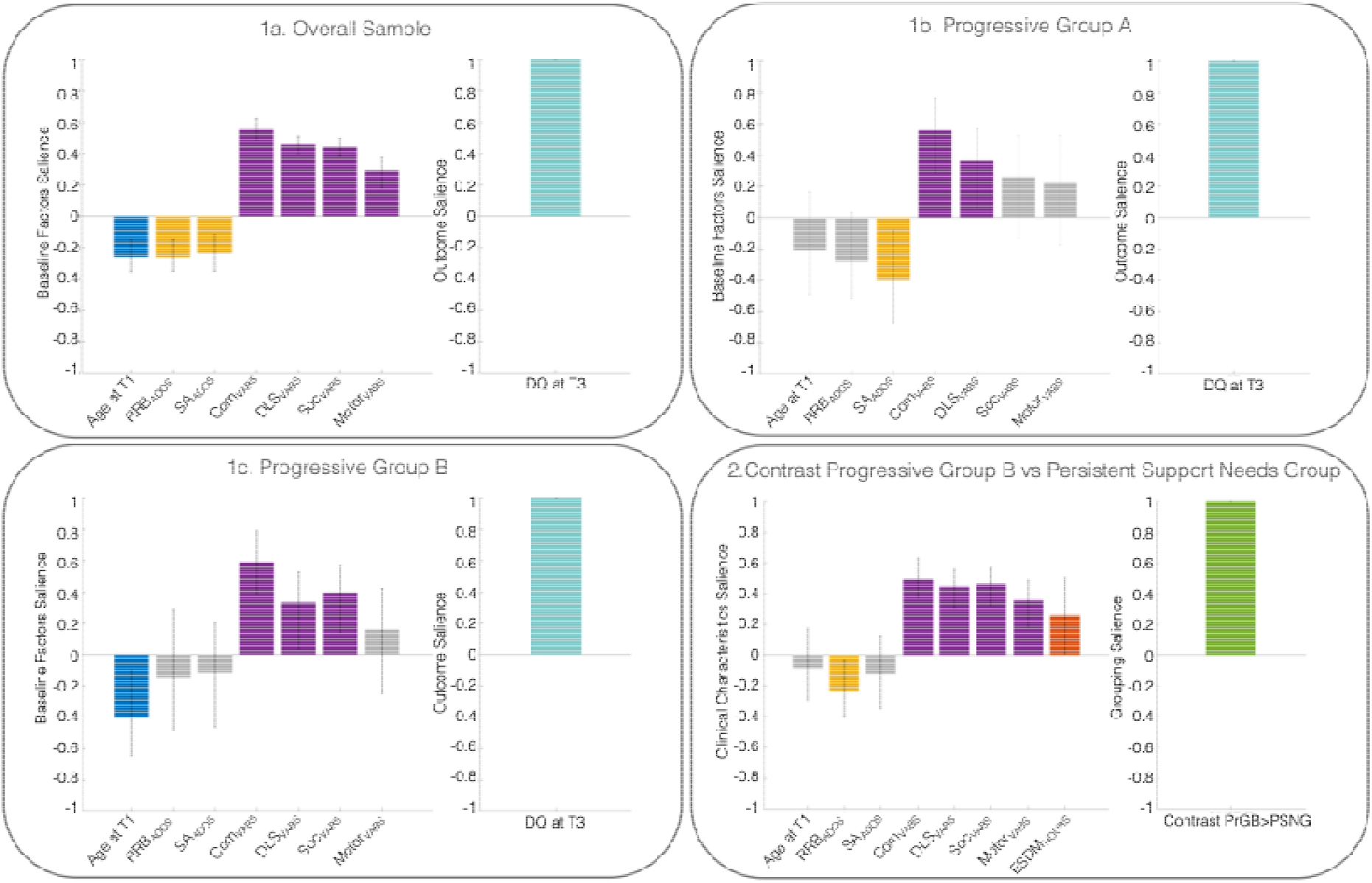
PLS-C of the baseline factors with the outcome across the original sample and the subgroups and contrast PLS between PrGB and PSNG. Notes. 3 In panels the bars on the left side of each panel represent the salience of the baseline variables contributing to the latent component associated with the outcome variable shown on the right side of the panel (DQ at T3 for panels 1a-1c and membership in Progressive Group B for panel 2). Colored bars indicate variables whose contribution to the latent component was stable across bootstrap resamples, whereas gray bars indicate variables that did not show stable contributions. The sign and magnitude of the salience values reflect the direction and strength of each variable’s contribution to the multivariate pattern associated with cognitive outcome. Panels 1a-c represent behavioral PLS-C and panel 2 represents contrast PLS-C between Progressive Group B and Persistent Support Needs Group. ADOS, Autism Diagnosis Observation Schedule; RRB_ADOS_; Restricted and Repetitive Behaviors, SA_ADOS_; Social Affect, VABS, Vineland Adaptive Behavior Scales – 2^nd^ Edition; Com_VABS_, Communication; DLS_VABS_, Daily Living Skills; Mot_VABS,_ Motor Skills; DQ, Developmental Quotient, ESDM_HOURS_; Total number of Early Start Denver Model intervention hours.

### 3.4 Predictors of the Cognitive Outcomes Within Subgroups

In addition to the latent pattern observed in the overall sample, we ran 3 separate PLS-C analyses for each subgroup to identify the baseline factors that were associated with the final cognitive outcome (Figure 3.1b, 3.1c). For the PrGA, one significant latent pattern (r = 0.57, p = 0.002) was identified including lower SA_ADOS_ and higher Com_VABS_ and DLS_VABS_ scores that were predictive of a higher cognitive outcome. Similarly, one significant latent pattern was identified for PrGB (r = 49, p = 0.004), where younger age at intake and higher baseline Com_VABS_, DLS_VABS_ and Soc_VABS_ were associated with a higher cognitive score by the end of the intervention. However, no significant latent component emerged within the PSNG.

### 3.5 Comparison between the Progressive Group B and the Persistent Support Needs Group

Finally, we wanted to better understand which clinical characteristics could distinguish PrGB from PSNG. At T1, both PrGB and PSNG exhibited average cognitive scores within the range of developmental delay. However, their developmental trajectories diverged substantially over the course of intervention. By T3, PrGB demonstrated the greatest cognitive gains among groups, with an average gain of 29.9 points of DQ, whereas PSNG showed substantially more limited progress and remained in the range of cognitive delay (mean DQ = 41.1). To further investigate the factors underlying the divergent developmental trajectories of PrGB and PSNG, we applied a contrast PLS-C analysis (Krishnan et al., 2011; McIntosh & Lobaugh, 2004) between these groups (Figure 3.2). One significant latent pattern was identified (r = 0.51, p = 0.001) indicating that higher adaptive skills, fewer RRBs at baseline and more hours of intervention were associated with membership in PrGB and distinguished this group from PSNG.

### 3.6 Comparison of the subgroups with the A3D Theoretical framework

In the last step of our analysis, we compared our three subgroups against the clustering solution provided by the A3D Theoretical framework. The comparison revealed good compatibility of the two-clustering models with 88.5% of the PSNG categorized as Type I and 85.6% of the PrGA and PrGB classified as Type II (Supplement C Table 1). This convergence suggests that PrGA and PrGB share developmental characteristics with the more typical advancement of Type II, whereas PSNG follows a slower developmental rate consistent with Type I.

## 4. DISCUSSION

This study examined the development of 125 autistic children receiving 12 to 20 weekly hours of ESDM intervention over a follow-up period of 1.93 years. On average children show a notable increase of 17 points in Composite Developmental Quotient (DQ) accompanied by significant improvement in their adaptive skills and reduction in autism-related features by the end of the intervention. Clustering revealed 3 distinct trajectories of response to intervention. The Progressive Group A (PrGA, 36%) exhibited the highest baseline DQ and achieved an average gain of 16.6 points. The Progressive Group B (PGrB, 41.6%) began with substantial developmental delay but showed the greatest average gain of 29.9 points, reaching the lower range of typical development by the end of the intervention. In contrast, the Persistent Support Needs Group (PSNG, 22.4%) presented the lowest baseline DQ and demonstrated a less steep developmental trajectory throughout the intervention. Using multivariate analysis, we identified distinct patterns of predictors associated with cognitive outcomes in the overall sample and within each subgroup. In PrGA, greater communication and daily living skills and lower levels of autism features in the domain of social affect at baseline were related to a higher cognitive outcome. In PrGB, younger age and higher adaptive skills in the domains of communication, socialization and daily living skills at intake were associated with better DQ scores by the end of intervention. However, no stable patterns of baseline factors related to the cognitive outcome were identified in PSNG. Finally, we performed a contrast PLS-C analysis comparing PrGB and PSNG, to disentangle the factors underlying their distinct developmental trajectories, despite both groups starting the intervention with cognitive skills in the range of developmental delay. Precisely, lower RRB_ADOS_ scores, higher adaptive skills at baseline and more hours of intervention were significantly associated with membership in PrGB group and differentiated this group from PSNG.

Our results align with previous research showing that the ESDM intervention is an effective method for the development of cognitive, adaptive skills and the reduction of autism features (Colombi et al., 2018; Mandelli et al., 2025; Wang et al., 2022). Our findings of 3 distinct developmental trajectories support prior evidence that the cognitive outcome is variable among autistic children, with the observed trajectories varying depending on the methodological approach applied (Asta et al., 2024; Godel et al., 2022; Lombardo et al., 2019; Mandelli et al., 2024). Indeed, a longitudinal study by Lord et al., (2015) following participants from 2 to 19 years old investigating the different autism phenotypes, proposed 3 distinct developmental trajectories based on their IQ, autism traits and adaptive functioning by the age of 19. Precisely, in that study the longitudinal trajectories of the participants represented two groups with great improvements across the years and a third group with fewer improvements and more stable development. Notably, the early developmental patterns reported in Lord et al., (2015) resemble the trajectories observed in our study during the overlapping developmental period. More recently, Mandelli and colleagues focused on the delineation of distinct subtypes of adaptive functioning, and identified 3 highly reproducible subgroups (high, medium and low) that were predictive of cognitive, verbal and non-verbal trajectories (Mandelli et al., 2023). We observe a high degree of overlap between our clusters and the ones obtained from A3D. Specifically, our two progressive groups share similar characteristics with Type II, representing a faster rate of developmental skills, while PSNG is more compatible with Type I demonstrating a less pronounced development over time. However, our approach expands the A3D framework by distinguishing two progressive groups rather than one group with favorable outcomes. This clustering highlights the heterogeneity in baseline cognitive levels capturing distinct developmental trajectories that provide a more detailed representation of responses to early intensive intervention. Consequently, differences in the number and characteristics of subgroups identified across studies likely reflect the inherent heterogeneity of autism, whereby developmental trajectories are shaped by diverse constellations of biological, cognitive, behavioral, and environmental factors. This underscores the importance of moving beyond average intervention effects toward approaches that characterize individual developmental pathways.

Similarly, while previous studies identified predictors by examining the overall sample, our approach additionally assesses subgroup specific patterns of predictors related with a higher cognitive outcome. Precisely, for PrGA, who start with a higher DQ, lower scores on social affect and greater communication and daily living skills were associated with a higher final cognitive outcome, whereas age at intake did not make a stable contribution to the predictive pattern. In contrast, for PrGB, children who entered the intervention with more pronounced developmental delays but ultimately achieved the greatest cognitive gains, younger age and stronger adaptive functioning across communication, socialization, and daily living domains were associated with better outcomes. Our results indicate that although adaptive skills are a common factor related to more favorable cognitive outcomes, the contribution of other baseline factors differs across subgroups. Specifically, lower scores on social affect are associated with better outcomes for children with higher cognitive skills at baseline, while younger age at the onset of the intervention is pivotal for the developmental progress of children with developmental delay. It is thus significant to consider developmental variability when identifying predictors of intervention response, as factors associated with favorable outcomes may differ depending on the child’s initial developmental profile rather than being homogeneously applicable across all autistic children. Our findings are consistent with prior evidence indicating that younger age is a robust predictor of better post-intervention outcomes (Devescovi et al., 2016; Mandelli et al., 2025; Vivanti et al., 2019). Similarly, Mandelli et al., (2023) showed that better adaptive functioning was predictive of more favorable non-verbal cognitive, language and fine motor scores in early childhood. Moreover, a recent meta-analysis has revealed that better cognitive and adaptive skills and lower scores on autism assessments were associated with better results from behavioral interventions (Chetcuti et al., 2025). Furthermore, communication skills at intake have been shown to play a pivotal role at the improvement of cognitive gain in early intensive intervention (Contaldo et al., 2020). A study examining the association of motor deficiencies with several autism features revealed that impaired fine and gross motor skills were associated with language difficulties and cognitive delay (Bhat et al., 2022). Additional analyses including the total number of intervention hours as well as demographic factors (family income, parental education, and parental age at the birth of the child) did not reveal stable associations with cognitive outcome at the end of the intervention. These findings were consistent across the full sample and within each response group.

Regarding the PSNG, which represented the smallest subgroup revealed by our clustering approach, children exhibited a developmental trajectory that differed markedly from those observed in the progressive responder groups. Although both PrGB and PSNG presented with cognitive scores in the delayed range at baseline (DQ < 70), children in PrGB achieved, on average, cognitive scores within the normative range at follow-up. On the other hand, children in PSNG remained in the delayed range. This finding should not be interpreted as a lack of improvement or a regression of skills. Because DQ is adjusted for chronological age, a stable DQ indicates that developmental gains occurred at approximately the same pace as chronological age increase, whereas increases in DQ reflect accelerated developmental progress. According to our results, none of the baseline factors included in the within-group PLS-C analysis reached statistical significance and thus we did not identify any stable patterns of predictors associated with a higher cognitive outcome for Persistent Support Needs Group. This likely reflects both the relatively small sample size and the greater homogeneity of developmental trajectories within this subgroup, which limit the ability of PLS-C to identify reliable covariance patterns between baseline characteristics and outcome.

Despite showing drastically different developmental trajectories, both the PSNG and PrGB group showed relatively similar developmental delays at the start of the intervention. To better understand the factors differentiating the trajectories of PSNG and PrGB, we conducted a contrast PLS-C analysis comparing the two groups. Thanks to the increased sample size, compared to our previous paper by Godel et al., (2022), we were able to uncover reliable patterns of predictors differentiating the two groups. Specifically, our results indicated that lower levels of RRBs, higher adaptive functioning at baseline, and a greater total number of ESDM intervention hours were associated with membership in PrGB and distinguished this group from PSNG. These findings suggest that among children with substantial developmental delay at the onset of the intervention, differences in adaptive functioning, autism features, and total number of intervention hours received contribute meaningfully to developmental outcomes and may help explain the divergence in trajectories observed between PrGB and PSNG. Our findings are partially consistent with those reported by Vivanti et al., (2025), who found that children who did not acquire word combinations by the end of early intervention were older at baseline and exhibited higher autism symptom severity as well as lower adaptive and cognitive skills than children who successfully acquired word combinations. Furthermore, the meta-analysis by Virués-Ortega (2010) highlighted the positive impact of greater intervention intensity on language and adaptive outcomes. Consequently, these findings suggest that autistic children with cognitive delay at the onset of intervention follow distinct developmental trajectories. Therefore, examining baseline factors that differentiate these trajectories allows us to move towards the idea of a “what works for whom” framework, with the goal of identifying individualized intervention approaches that may optimize developmental outcomes for children presenting with lower baseline functioning and more persistent support needs. Notably, the factors distinguishing developmental trajectories in the present study included both child-related characteristics (e.g., adaptive functioning and symptom profile) and intervention-related factors (e.g., cumulative intervention exposure). Interestingly, neither age at intake nor socioeconomic factors differentiated the response groups, suggesting that the mechanisms underlying variability in treatment response are likely more complex than simple differences in treatment access or timing of intervention. Future research should therefore adopt a more holistic perspective that considers not only the child’s developmental profile, but also intervention characteristics and broader family-related factors, in order to better understand the mechanisms underlying individual differences in intervention response. Equally important is the consideration of a broader range of outcomes, including language, symptom levels, cognitive and adaptive functioning to capture progress in different developmental levels and enhance our understanding on response patterns to early intensive intervention (Zwaigenbaum et al., 2015). Such efforts are essential for moving beyond average treatment effects and toward precision intervention models.

Although this study includes, to our knowledge, the largest monocentric sample of autistic preschoolers receiving ESDM intervention over a 2-year follow-up period, stratification of the original sample into distinct developmental trajectories resulted in substantially smaller subgroup sizes and especially for the PSNG. Consequently, the limited number of participants in PSNG did not allow the identification of a stable pattern of factors associated with the final cognitive outcome. Another limitation of the present study is the use of different cognitive evaluation tools namely MSEL, PEP-3 and WPPSI-IV. The inclusion of participants assessed with PEP-3 and WPPSI-IV allowed us to increase the sample size, thereby enhancing the statistical power and generalizability of the findings. While there was a strong positive correlation between the MSEL and PEP-3, these assessments differ in their structure and therefore cannot be considered fully equivalent. Although these participants represent a small part of our sample (PEP-3, n = 28 ; WPPSI-IV, n = 2), this variance in cognitive measures should be considered with great caution when interpreting the results. Finally, the study design did not allow the comparison of ESDM intervention with alternative intervention approaches or community-based services. Consequently, while we observed significant developmental gains among children receiving ESDM intervention, future RCTs are needed to define to what extent this progress can be attributed to ESDM.

In conclusion, parsing the heterogeneity of response to ESDM intervention by identifying distinct response subgroups and modeling subgroup-specific predictors moves beyond the assumption that the same factors influence all children’s cognitive outcome. This approach enhances our understanding of the unique characteristics of the response groups and the mechanisms driving this heterogeneity, providing the opportunity for more personalized recommendations for intervention from clinicians.

## Supporting information

## Data Availability

All data produced in the present study are available upon reasonable request to the authors.

## Acknowledgments

The authors would like to thank all the families that participated in this research, all the therapists at the Centres d’Intervention Précoce en Autisme of the Fondation Pôle Autisme (https://pole-autisme.ch) and the Office Médico-Pédagogique in Geneva. In addition, the authors would like to thank Margot Giraud, Niveettha Thillainathan, Pamela Iraci, Stefania Solazzo, Michel Godel and numerous other lab members who contributed to data acquisition over the years.

This research was conducted thanks to funding to I.I. through the NCCR Evolving Language, Swiss National Science Foundation Agreement #51NF40_225146. The data acquisition over the years that has made this study possible were supported by several funding sources: the Swiss National Foundation Synapsy (Grant No. 51NF40–185897), the Swiss National Foundation for Scientific Research (Grant Nos. #163859, #190084, #202235, #212653 to M.S. and Ambizione Grant n° PZ00-1_233424 / 1 to N.K.), the Fondation Privée des Hôpitaux Universitaires de Genève (https://www.fondationhug.org) and by the Fondation Pôle Autisme (https://www.pole-autisme.ch). The funders provided only financial support and were not involved in the study design.

## Notes

**Conflict of Interest:** The authors declare no conflict of interest. The funders did not participate in the design of the study, the collection, the analysis of the data, or the interpretation of the results.

### Competing Interest Statement

The authors have declared no competing interest.

### Author Declarations

The Regional Research Ethics Committee (CCER) (Protocol Number: 2025-00009) gave ethical approval for this work.

### Summary of Updates

In response to the reviewers' comments, we substantially revised the manuscript by increasing the sample size from 107 to 125 participants, refining the clustering and statistical approach. These changes strengthen the methodological rigor, interpretability, and clinical relevance of the study.

