## Supplementary material for "Heterogeneity of response to Early Start Denver Model: Identifying developmental trajectories and predictors of cognitive outcomes"

**SUPPORTING INFORMATION**

**SUPPLEMENT A: METHODS**

**ESDM Intervention**

The 125 participants of our study received 12-20h per week of ESDM intervention. For the participants that the end of the intervention occurred between two scheduled follow-up visits, we wanted to ensure a consistent and objective selection of the outcome timepoint to guarantee that the end of the intervention is close to the outcome timepoint. Precisely, we used the first visit following the end of the intervention as outcome timepoint, unless the preceding visit occurred within one month prior to the end of the intervention, in which case the earlier visit was retained. For 95 participants the outcome timepoint was the 2 years follow-up and for 30 participants it was the 1.5 year follow-up visit. Among the 30 participants with 1.5 year of follow-up as outcome timepoint, 9 participants are currently enrolled in the ESDM intervention program and do not have a 2 years follow-up yet. Sixteen participants have a 2 year follow-up but the 1.5 year follow-up was selected as an outcome timepoint for the reasons explained above. Finally for 5 participants the 2 years follow-up was not available due to early drop-out from the intervention program or unavailability of the family for the last follow-up.

The therapists that provide ESDM intervention received training during the CIPA program according to the ESDM Teaching Fidelity Rating System (Rogers & Dawson, 2010). The team consists of Master-level qualified therapists that are supervised by a certified trainer. Even though the intervention is offered in different departments, there are no variances in the ESDM instruction, fidelity rating assessment and guidance by the certified trainer.

**Cognitive evaluations and computation of Composite DQ**

Cognition during childhood has been found to be a strong predictor of future outcome during adulthood including later school and academic achievements but also independence (Billstedt et al., 2005; Gillespie-Lynch et al., 2012; Howlin et al., 2004). Cognitive skills are the domain that is most frequently reported in previous intervention studies but also the domain that shows the biggest progress in children receiving early intensive intervention. This is why we decided to use DQ as the main outcome variable in our study (Godel et al., 2022; Rogers et al., 2019; Rogers & Dawson, 2010). The domains of adaptive skills and autism features have also been selected as secondary outcomes to better capture the heterogenous developmental trajectories.

As mentioned above all children were evaluated with one or more of the following cognitive assessments: MSEL, PEP-3 and WPPSI-IV. The MSEL includes the subdomains of Gross Motor (GM), Visual Reception (VR), Fine Motor (FM), Expressive Language (EL) and Receptive Language (RL) (Mullen, 1995). PEP-3 includes verbal and preverbal cognition (VPC), Expressive Language (EL), Receptive Language (RL), Fine Motor (FM), Gross Motor (GM), Visuo-Motor Imitation (VMI) scales (Schopler et al., 2005). WPPSI-IV provides a Full Scale Intellectual Quotient (FSIQ) as well as 5 primary index scores; Verbal Comprehension (VC), Visual Spatial (VS), Fluid Reasoning (FR), Working Memory (WM), and Processing Speed (PS) (Salonen et al., 2023; Wechsler, 2012). Since the various cognitive tests differ in structure and provide different scores, we aimed to compute a Composite DQ that provides a common measure of the overall cognition. To calculate the Composite DQ from MSEL we averaged the developmental ages of: VR, FM RL, and EL, then we divided by chronological age and multiplied by 100 (Godel et al., 2022; Lord et al., 2006). We repeated the same process for PEP-3 using the subdomains of: VPC, EL, RL, and FM, as described in 2022 by Godel et al. For the 2 cases that WPPSI-IV was used as a cognitive assessment, we considered the Intellectual Quotient of the test as equivalent of Composite DQ.

To estimate the coherence of MSEL and PEP-3, we examined 92 participants at T1 that were evaluated with both evaluations. According to the results there was a strong positive correlation between the composite scores of MSEL and PEP-3, r(90) = 0.842, p < 0.001.

**Supplement B: RESULTS**

**General Trajectory of the sample**

Table 1 Comparison of the evolution of the sample across the 3 timepoints.

| **Measures** | **p-value (Effect size)** | | |
| --- | --- | --- | --- |
|  | **T2-T1** | **T3-T1** | **T2-T3** |
| **Composite DQ** | <0.001 (-0.71) | <0.001 (-0.70) | ns |
| **ADOS-2** | <0.001 (-0.51) | <0.001 (-0.35) | 0.035 (0.22) |
| **SA (ADOS-2)** | <0.001 (-0.57) | <0.001 (-0.43) | ns |
| **RRB (ADOS-2)** | <0.001 (-0.32) | 0.014 (-0.25) | ns |
| **VABS-II** | <0.001 (-0.32) | <0.001 (-0.45) | 0.001 (-0.30) |

Notes. 1 p-values and effect sizes of Wilcoxon Signed Ranks Test for the evaluation of the sample throughout the 3 timepoints (T1,T2,T3) of ESDM intervention. FDR applied for multiple comparisons. The effect sizes are computed using the formula r=Z/√N introduced by Rosenthal, 1991, where Z is the Z-score and N is the number of observations.

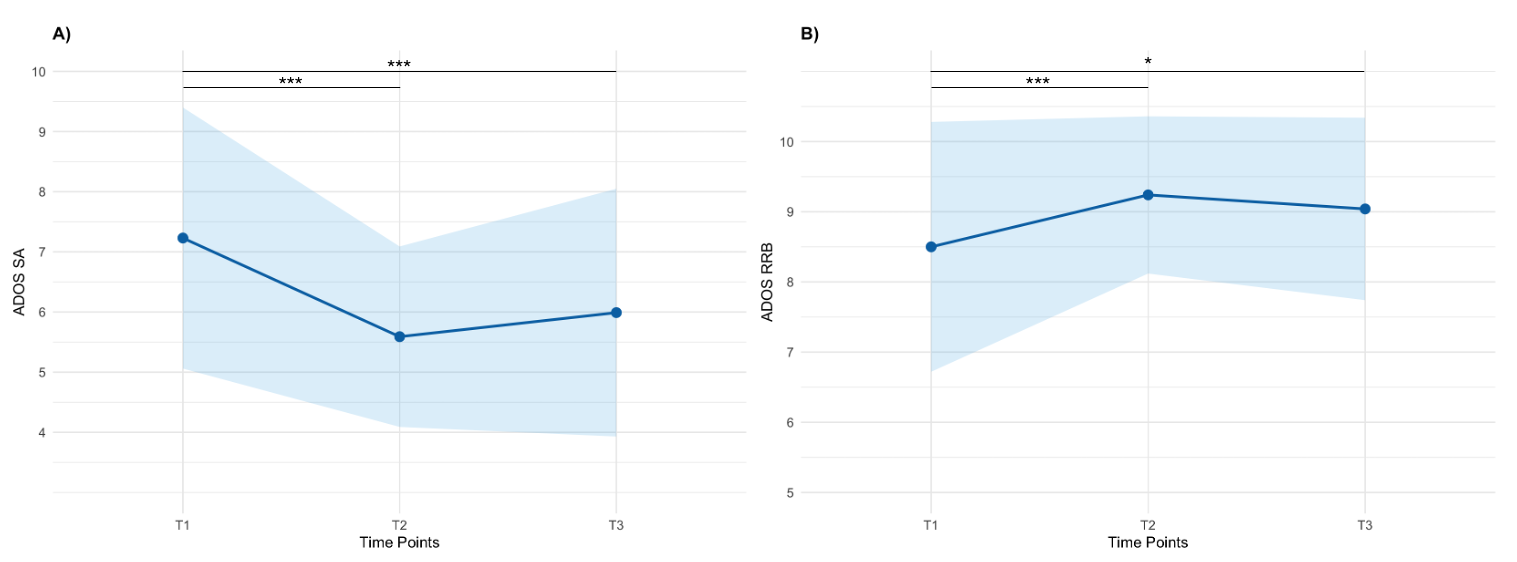

Notes.4 Developmental trajectories of participants across: A) SA_ADOS_, B) RRB_ADOS_. Blue bands signifying Standard Deviation. Pairwise comparisons were conducted using Wilcoxon Signed Ranks Test. FDR applied for multiple comparisons. Each line chart was created including the participants without missing values across all three timepoints (n=92). The corresponding p-values were computed including participants without missing values between the timepoints of comparison. Scores with a significant statistical difference are highlighted in bold; *p < 0.05, **p < 0.01, ***p < 0.001. ADOS, Autism Diagnosis Observation Schedule; SA, Social Affect, RRB, Restricted and Repetitive Behaviors

Figure 1 Longitudinal trajectories of the Overall Sample in the domains of Social SA_ADOS_ and RRB_ADOS_

Table 2 Comparison of the 3 subgroups across different measures

| **Measures** | **p-Value (Z score)** | **Effect Size** |
| --- | --- | --- |
| **RRB_ADOS_** | | |
| PrGA - PSNG | <0.001 (-3.580) | -0.42 |
| PrGA-PrGB | 0.044 (-2.019) | -0.20 |
| PSNG - PrGB | 0.034 (-2.119) | -0.24 |
| **SA_ADOS_** | | |
| PrGA - PSNG | 0.005 (-2.817) | -0.33 |
| PrGA-PrGB | 0.028 (-2.194) | -0.22 |
| PSNG - PrGB | ns | - |
| **Age** | | |
| PrGA - PSNG | 0.003 (-2.978) | -0.35 |
| PrGA-PrGB | 0.011 (-2.543) | -0.26 |
| PSNG - PrGB | ns | - |
| **DQ** | | |
| PrGA - PSNG | <0.001 (-7.113) | -0.83 |
| PrGA-PrGB | <0.001 (-8.319) | -0.84 |
| PSNG - PrGB | <0.001 (-3.389) | -0.38 |
| **SPC** | | |
| PrGA - PSNG | <0.001 (-5.877) | -0.69 |
| PrGA-PrGB | <0.001 (-6.547) | -0.66 |
| PSNG - PrGB | <0.001 (-7.293) | -0.82 |
| **Communication _VABS_** | | |
| PrGA - PSNG | <0.001 (-6.831) | -0.80 |
| PrGA-PrGB | <0.001 (-6.217) | -0.63 |
| PSNG - PrGB | <0.001 (-4.351) | -0.49 |
| **Daily Living Skills _VABS_** | | |
| PrGA - PSNG | <0.001(-5.739) | -0.67 |
| PrGA-PrGB | <0.001(-3.513) | -0.36 |
| PSNG - PrGB | <0.001(-3.608) | -0.40 |
| **Socialization _VABS_** | | |
| PrGA - PSNG | <0.001(-5.75) | -0.67 |
| PrGA-PrGB | <0.001(-3.854) | -0.39 |
| PSNG - PrGB | <0.001(-3.845) | -0.43 |
| **Motor Skills _VABS_** | | |
| PrGA - PSNG | <0.001(-3.873) | -0.45 |
| PrGA-PrGB | ns | - |
| PSNG - PrGB | 0.001(-3.263) | -0.36 |

Notes. 2. p-values computed using non-parametric Mann-Whitney-U Tests for the comparison between the groups. The effect sizes are computed using the formula r=Z/√N introduced by Rosenthal, 1991, where Z is the Z-score and N is the number of observations. PrGA, Progressive Group A; PrGB, Progressive Group B; PSNG, Persistent Support Needs Group.

**Supplement C**

Table 1 Comparison of our clustering with A3D Theoretical framework

|  | **TYPE I** | **TYPE II** |
| --- | --- | --- |
| **PrGA** | 0 | 45 |
| **PrGB** | 14 | 38 |
| **PSNG** | 23 | 3 |

Notes. 1 PrGA, Progressive Group A; PrGB, Progressive Group B; PSNG, Persistent Support Needs Group.
